# Delineating the heterogeneity of preimplantation development via unsupervised clustering of embryo candidates for transfer using automated, accurate and standardized morphokinetic annotation

**DOI:** 10.1101/2022.03.29.22273137

**Authors:** Nir Zabari, Yoav Kan-Tor, Yuval Or, Zeev Shoham, Yoel Shofaro, Dganit Richter, Iris Har-Vardi, Assaf Ben-Meir, Naama Srebnik, Amnon Buxboim

## Abstract

The majority of human embryos, whether naturally or *in vitro* fertilized (IVF), do not poses the capacity to implant within the uterus and reach live birth. Hence, selecting the embryos with the highest developmental potential to implant is imperative for improving pregnancy rates without prolonging time to pregnancy. The developmental potential of embryos can be assessed based on temporal profiling of the discrete morphokinetic events of preimplantation development. However, manual morphokinetic annotation introduces intra- and inter-observer variation and is time-consuming. Using a large clinically-labeled multicenter dataset of video recordings of preimplantation embryo development by time-lapse incubators, we trained a convolutional neural network and developed a classifier that performs fully automated, robust, and standardized annotation of the morphokinetic events with R-square 0.994 accuracy. To delineate the morphokinetic heterogeneity of preimplantation development, we performed unsupervised clustering of high-quality embryo candidates for transfer, which was independent of maternal age and blastulation rate. Retrospective comparative analysis of transfer versus implantation rates reveals differences between embryo clusters that are distinctively marked by poor synchronization of the third meiotic cell-cleavage cycle. We expect this work to advance the integration of morphokinetic-based decision support tools in IVF treatments and deepen our understanding of preimplantation heterogeneity.

## 1. Introduction

Owing to the inherent biological heterogeneity in the developmental potential of embryos, decreasing the risks that are associated with multiple pregnancy while shortening time to pregnancy relies on transferring the embryo(s) of the highest developmental quality.^1^ To address this important need, various assisted reproductive technologies (ARTs) have been developed.^1^ Embryos that are generated via *In Vitro* Fertilization (IVF) and harbor chromosomal abnormalities that are expected to decrease the potential to implant and generate a live birth can be screened via preimplantation genetic testing for aneuploidy (PGT-A) and for structural chromosomal rearrangements (PGT-SR).^2^ Recent advances in shifting towards genome wide single nucleotide polymorphism (SNP) microarrays and next generation senescing (NGS) allowed combining different PGT approaches for screening of chromosomal and genetic disorders.^3-7^ However, PGT is invasive and requires obtaining cellular biopsies from the embryos. In addition, false negative assessments may arise due to insufficient genetic amplification and chromosomal mosaicism.^8,9^

Complementary to PGT, various non-invasive technologies have been developed during the past decade for assessing embryo quality and developmental potential in real time and non-invasively.^10^ These approaches are based on the assumptions that embryo quality can be linked with certain metabolic markers, which can be assessed by probing molecular signatures in the spent culture medium.^11-14^ Other approaches relay on the mechanical changes in the viscoelastic properties of preimplantation embryos that are associated with high developmental quality and can be defined by measuring the stress-strain relationships under applied load.^15^ However, the most reliable methods for predicting embryo quality were based on visualization of specific developmental stages that can be scored using morphological grading criteria.^16-19^

The utilization of time-lapse incubation systems in IVF clinics provided dynamic and continuous visualization of the embryos as they monotonically advance between the states of preimplantation developmental while maintaining the embryos under optimal culture conditions (Fig 1). Using these video recordings, the embryos can be characterized by the specific time points from fertilization at which discrete developmental events occur. These so-called morphokinetic events are defined by the transition between embryo states and include times of pronuclei appearance (tPNa) and fading (tPNf), the cleavage of two-to-eight blastomeres (tN, N=1 to 8), the compaction of the morula (tM), and start of blastulation (tSB).^19,20^ The generation of large datasets of morphokinetically annotated and clinically labeled embryos facilitated the development of classification algorithms that predict the potential for embryo implantation^21,22^ and live birth.^23-25^ Parallel efforts took advantage of the size of the available time-lapse datasets to train convolutional neural network (CNN) based classifiers that assess embryo potential using the raw video files in an unbiased annotation-independent manner.^26,27^ However, training such deep learning models is challenging due to the size of the video files (∼100’s Mb), which would require a sufficiently large sample number.^28,29^

**Fig 1.**
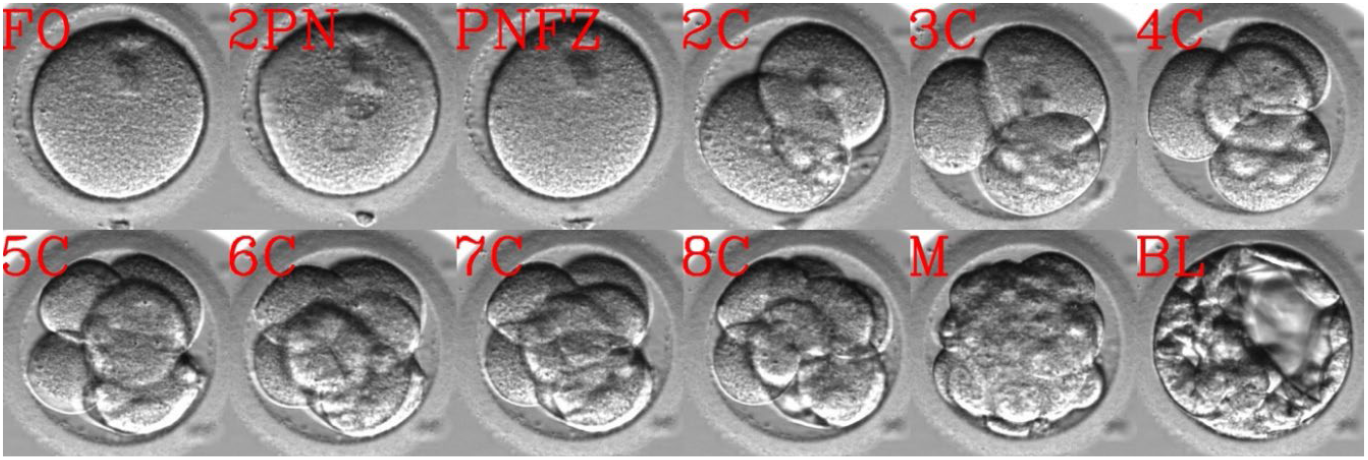
Preimplantation embryo development. Time lapse visualization of the developmental states of an embryo are demonstrated during preimplantation development. Abbreviations. FO: Fertilized oocyte. 2PN: Two pronuclei. PNFZ: Pronuclei faded zygote. 2C – to – 8C: Two – to – eight cells. M: Morula. BL: Blastocyst.

Morphokinetic evaluation of the developmental potential proved highly efficient in de-selecting for transfer poor quality embryos. However, annotation by trained embryologists of all the embryos from each oocyte collection cycle is time consuming. Moreover, manual morphokinetic annotation can introduce inter-observer and intra-observer variations. To support automatic and standardized morphokinetic assessment of embryo developmental potential, we used a large dataset of manually-annotated and clinically-labeled embryo video files, and trained a CNN classifier on the individual embryo frames to infer the extracted developmental states. By allowing the prediction of a superposition of multiple states at different probabilities, we obtained access to the hidden information that is stored by the morphokinetic uncertainties. We then constrained chronological development as appears by the time-lapse frame-wise series of developmental states to obtain the morphokinetic profiles of the embryos. Indeed, we predict the developmental states of the single frames with 97% accuracy and the series of morphokinetic profiles of the embryos from time of pronuclei appearance to start of blastulation with R-squared coefficient of determination 0.994 as validated across 1918 test set embryos. Using our automatic classifier, we provide unparalleled temporal statistics of preimplantation development of 67707 embryos. Focusing on 14159 high-quality embryos that would correspond to valid candidates for transfer, we apply unsupervised clustering into nine distinctive cohorts. We define distinctive patterns of early and late morphokinetics and reveal cluster-specific correlations with the rate of embryo transfer and the rate of embryo implantation. Our work thus provides a standardized platform for assessing the developmental potential of pre-implantation embryos. By supporting single embryo transfer policies, our work is expected to decrease the medical risks that are associated with multiple pregnancies and shorten time to pregnancy.

## 2. Methods

### 2.1. Dataset

The extraction of morphokientic profiles from embryo time-lapse imaging data is a challenging task owing to the chronological nature of preimplantation development and the dimensionality of the video files. To develop an automatic, accurate and standardized algorithm for extracting the morphokinetic events, we used a previously assembled database.^27^ In short, we assembled a large dataset of 67,707 video files of preimplantation embryo development that were recorded on eleven time-lapse incubation systems (Embryoscope, Vitrolife) located in four medical centers. The dataset includes 20,253 embryos that were morphokinetically manually-annotated by trained embryologists adhering to established protocols as we reported previously.^27^ Time-lapse images were recorded with an average 18 min time interval for 3-to-6 days. At each time point, seven Z-stack frames were recorded, however only the central focal plane was used here. The embryos in the dataset were randomly divided into a train-set (80%) and a test-set (20%) such that frames of individual embryos were not shared between them (Table-1). The train-set was further divided by randomly removing 15% of the frames to the validation set. Unlike the validation and test sets, which include an unbiased distribution of transferred and non-transferred embryos, only transferred embryos were actually considered for network training.

**Table-1:** Number of test-set frames across embryo states.

| Embryo state | 1C | PN | 2C | 3C | 4C | 5C | 6C | 7C | $\geq 8C$ | M | $\geq SB$ |
| --- | --- | --- | --- | --- | --- | --- | --- | --- | --- | --- | --- |
| Train set frames | 67498 | 174598 | 103955 | 15744 | 105914 | 27651 | 26943 | 31671 | 241313 | 107470 | 244581 |
| Test set frames | 27132 | 70428 | 42738 | 6083 | 42832 | 10581 | 10949 | 11437 | 98698 | 42611 | 101814 |

### 2.1. Embryo-state frame labeling

The morphokientic events characterize the preimplantation dynamics of the embryos and are not a property of the individual time-lapse frames. Hence, we first converted the manually-annotated morphokinetic profiles of the embryos into the so-called developmental embryo state labels of each individual frame. Given the monotonic nature of preimplantation development, the conversion of the manually annotated morphokinetic profiles into embryo state labels was performed in straightforward manner as specified in Table-2. Frames that overlap the manually annotated morphokientic events and the frames that were recorded just after were excluded from the train set. Notably, here we discriminate between *FO* and *PNFZ* embryo states despite being morphologically-identical. Hence, *FO* and *PNFZ* frames were re-labeled one cell (1*C*) for the purpose of network training.

**Table-2:** Rules for setting the embryo states of the frames based on the profiles of morphokinetic events. FO: fertilized oocyte. *tPNa* and *tPNf*: time of PN appearance and fading. *t*_*N*_: time of N blastomeres cleavage event. *t*_*M*_: time of Morula compaction. *t*_*SB*_: time of start-of-blastulation. PN: pronuclei. PNFZ: PN-fading zygote.

| Morphokinetic event | Embryo state |
| --- | --- |
| $t < t_{PNa}$ | *FO* |
| $t_{PNa} \leq t < t_{PNf}$ | *PN* |
| $t_{PNf} \leq t < t_2$ | *PNFZ* |
| $t_N \leq t < t_{N+1}$ | *NC*, $2 \leq N \leq 7$ |
| $t_8 \leq t < t_M$ | $8C^+$ |
| $t_M \leq t < t_{SB}$ | *M* |
| $t_{SB} \leq t$ | $SB^+$ |
| $t = t_n$ or $t_{n+1}$ | Frames discarded from train-set |

### 2.2. Frame preprocessing

The Embryoscope time-lapse incubator (Vitrolife) records 8-bit grayscale images that are composed of 500×500 grayscale pixels. To decrease dimensionality, a 256×256 pixels region of interest (ROI) of the embryos in each frame was cropped using a U-net segmentation network as we reported previously.^27^ In addition, all train-set frames were further augmented by applying [90°, 180°, 270°] rigid rotations, horizontal flipping, and vertical flipping.

### 2.3. Inference of the frame-wise embryo-state probability vector and the embryo probability matrix

To infer the developmental states of the embryo as visualized in each individual frame, we trained a ResNet18 CNN,^30^ using the train, validation and test sets of the time-lapse frames labeled by the embryo developmental states as described above. Since these are grayscale image files, we modified the first convolutional layer to input a single channel instead of three. The CNN model was implemented using TorchVision in PyTorch with a categorical cross entropy loss function, and optimized using Rectified Adam (RAdam). A learning rate was set to 0.0005, reaching convergence within ten epochs.

Let us consider the time-lapse sequence of size *n* of a given embryo, where *t*_*i*_ is the time from fertilization of frame *i* = 1, …, *n*. The classifier infers the probability to find the embryo at any of the developmental states, from 1C to BL, as obtained by the eleven output neurons (Fig 2). In this manner, the weighted and superimposed developmental state of frame *i* is defined by the embryo state probability vector (ESPV) of the output neuron coordinates:

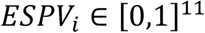

**Fig 2.**
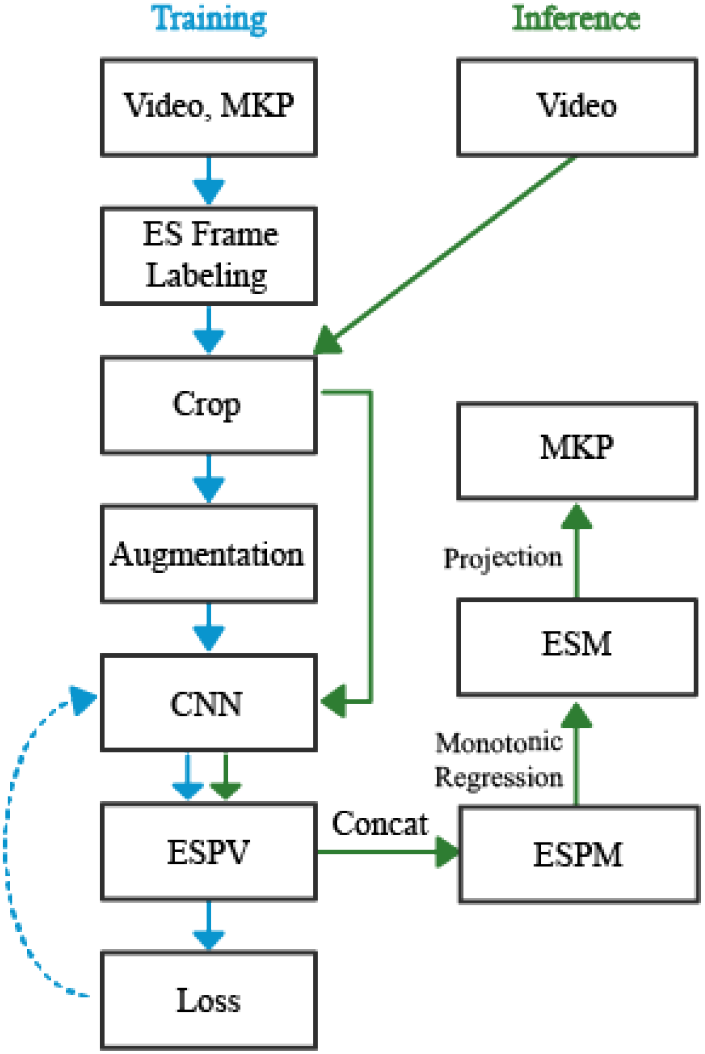
Training a CNN model for performing automated morphokinetic annotation of embryo video files. Training (blue): The classifier is trained using manually-annotated time-lapse images of preimplantation embryos. Based on the manually-labeled profiles the morphokinetic events (MKPs) of the videos, the temporal embryo states are extracted, which are then used as labels of each individual frame. Following a standard preprocessing step (cropping and augmentation of the frames), a CNN is trained for predicting the ESPV using a cross entropy loss function. Inference (green): An ESPV is generated for each frame in the video by the trained CNN model. For each embryo, the ESPM is generated via chronological concatenation of the ESPVs, which is then projected onto the ESM via monotonic regression. Temporal discrete transitions in the embryos sates as depicted by the ESM are converted into a MKP of the embryo. CNN: convolutional neural network. ESPV: embryo state probability vector. ESPM: embryo state probability matrix. ESM: embryo state matrix. MKP: morphokinetic profile.

The ESPVs of five representative snapshots at ascending order are shown in Fig 3A. To obtain a probability-weighted whole-embryo dynamic representation of preimplantation development, we define the embryo-state probability matrix (ESPM) by concatenating all the ESPVs of the embryo in a chronological order (Fig 3B-i).

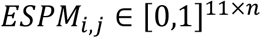

**Fig 3.**
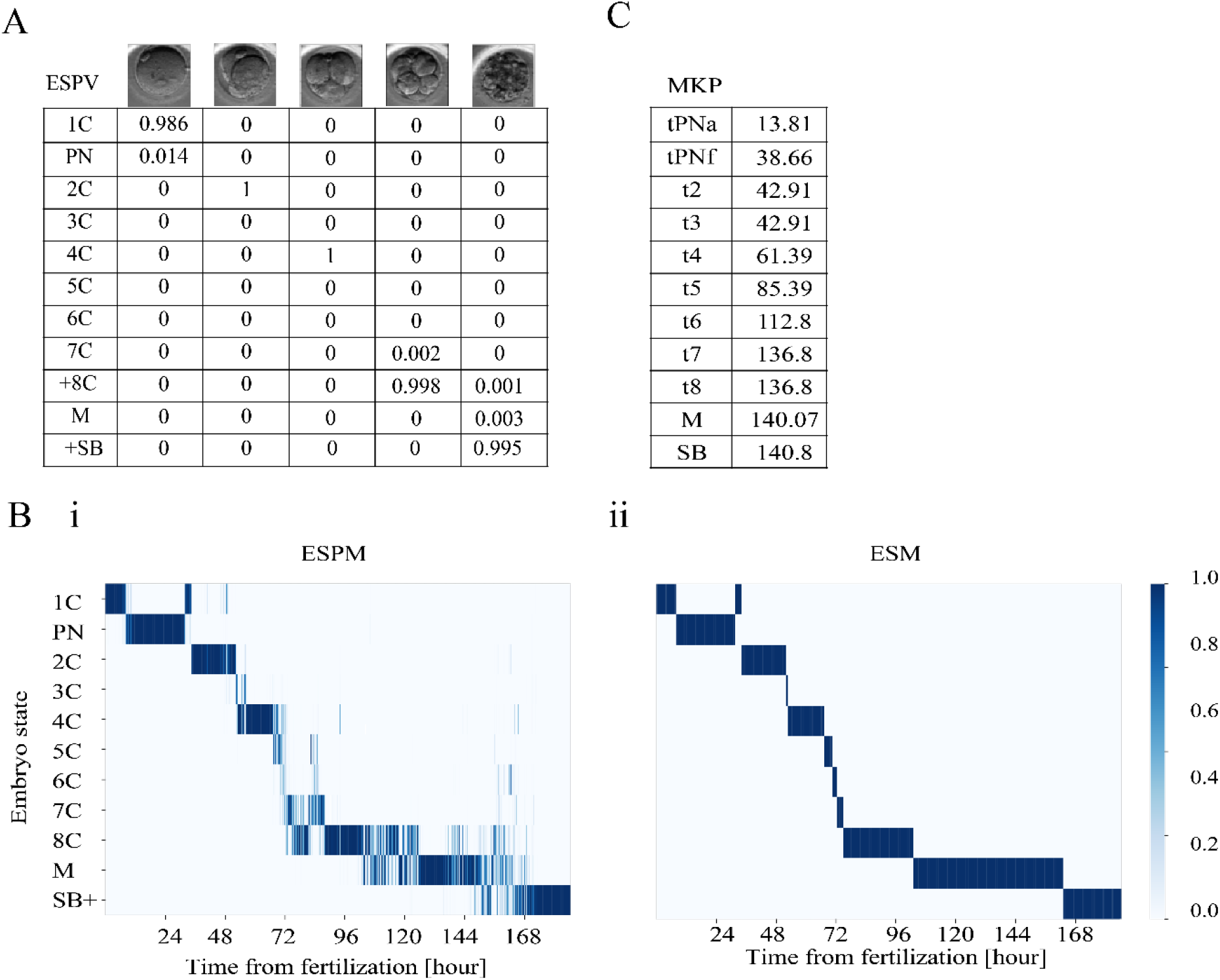
Automated morphokinetic annotation of embryo preimplantation development from ESPV to MKP. Automated morphokientic annotation is demonstrated for a representative embryo. (**A)** The ESPM of the embryos is generated via chronological concatenation (left to right) of the ESPV columns. (**B)** (i) A heatmap presentation of the ESPM superimposes multi-state distributions of the embryo at each time point (columns). (ii) Monotonic regression is used to project the superimposed states onto a single state at each time point as appear by the ESM. The FO and the PNFZ, which appear just before and right after the 2PN state, share a one-cell (1C) label owing to their indistinguishable morphology. (**C)** The MKP are set by discrete events of the temporal transitions between embryo states in the ESM. MKP: Morphokinetic profiles. ESPV: embryo state probability vector. ESPM: embryo state probability matrix. ESM: embryo state matrix. FO: Fertilized oocyte. PNFZ: Pronuclei faded zygote states. 2PN: Two pronuclei.

### 2.4. Automatic annotation of the morphokientic events

To extract the discrete time points of the morphokinetic events, the uncertainty in the developmental states of the embryo, as formulated by the ESPM, should first be projected onto discrete temporal states. Hence, we project ESPM onto 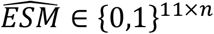 as follows:

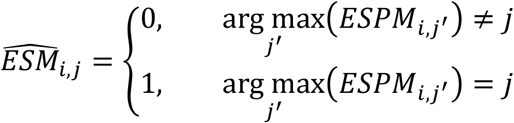

While 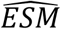 provides a discrete description of the temporal states of the embryo, it does not satisfy the monotonicity of preimplantation development. To this end, we apply a weightless isotonic regression of 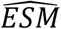 using the scikit-learn package in Python:

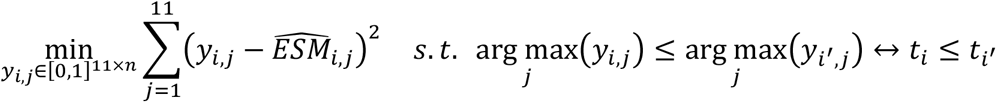

Next, we obtain the binary and discrete matrix *ŷ* ∈ [0,1]^11×*n*^ as follows (Fig 3B-ii):

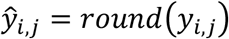

We recall that FO and PNFZ are two embryo states that share the 1C morphological label which is used here. Hence, we expect to obtain at least two temporally-separated regions in the ESPM of value 1C (see Fig 3B-i) that will be propagated to 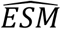 but converged in *ŷ*. Based on trial-and-error, we find that FO and PNFZ are best captured by the earliest and the latest 1C regions in *ŷ*, respectively. Hence, we replace the FO and PNFZ time regions (first row) in *ŷ* and obtain the binary and developmentally-monotonic embryo state matrix *ESM* ∈ {0,1}^11×*n*^ (Fig 3B-ii).

With increasing *i* (*ESM* columns), there are up to eleven transitions between the embryo states that correspond to the morphokinetic events of that embryo, which we extract in a straightforward manner. The time of the morphokinetic event *i* is thus defined by the transition from embryo state *j* to the consecutive one (most frequently *j* + 1). In the case of direct equal cleavage from *m* cells, *m* > 1, to (*m* + 2) cells, the morphokinetic events (*m* + 1)*C* and (*m* + 2)*C* will converge. The vector of time points of the morphokientic events is the automatically annotated morphokinetic profile of that embryo (Fig 3C).

## 3. Results

### 3.1. Inference uncertainty of the frame-wise embryo states

Using a probabilistic presentation of the embryo states, as presented by the ESPM, we propagate the information that is stored by the uncertainty in each frame (Fig 4A – top panels). We find that the regions of high uncertainty change between embryos, however noise tends to be high during the second (3C to 4C) and third (4C to 8C) embryo cleavage blocks and during morula compaction (Fig 4B). In the process of projection of the ESPM onto single embryo states, as presented by the (ESM), we take into account the temporal neighborhood constrains of the temporal monotonicity of preimplantation development (Fig 4A – bottom panels and Fig 4B). Indeed, morphokinetic annotation tends to either be in agreement with manual annotation, or to be separated by one or two sequences (Fig 5). Given the discrete nature of time-lapse imaging, the basal sampling error for morphokinetic annotation is set by one interval difference.

**Fig 4.**
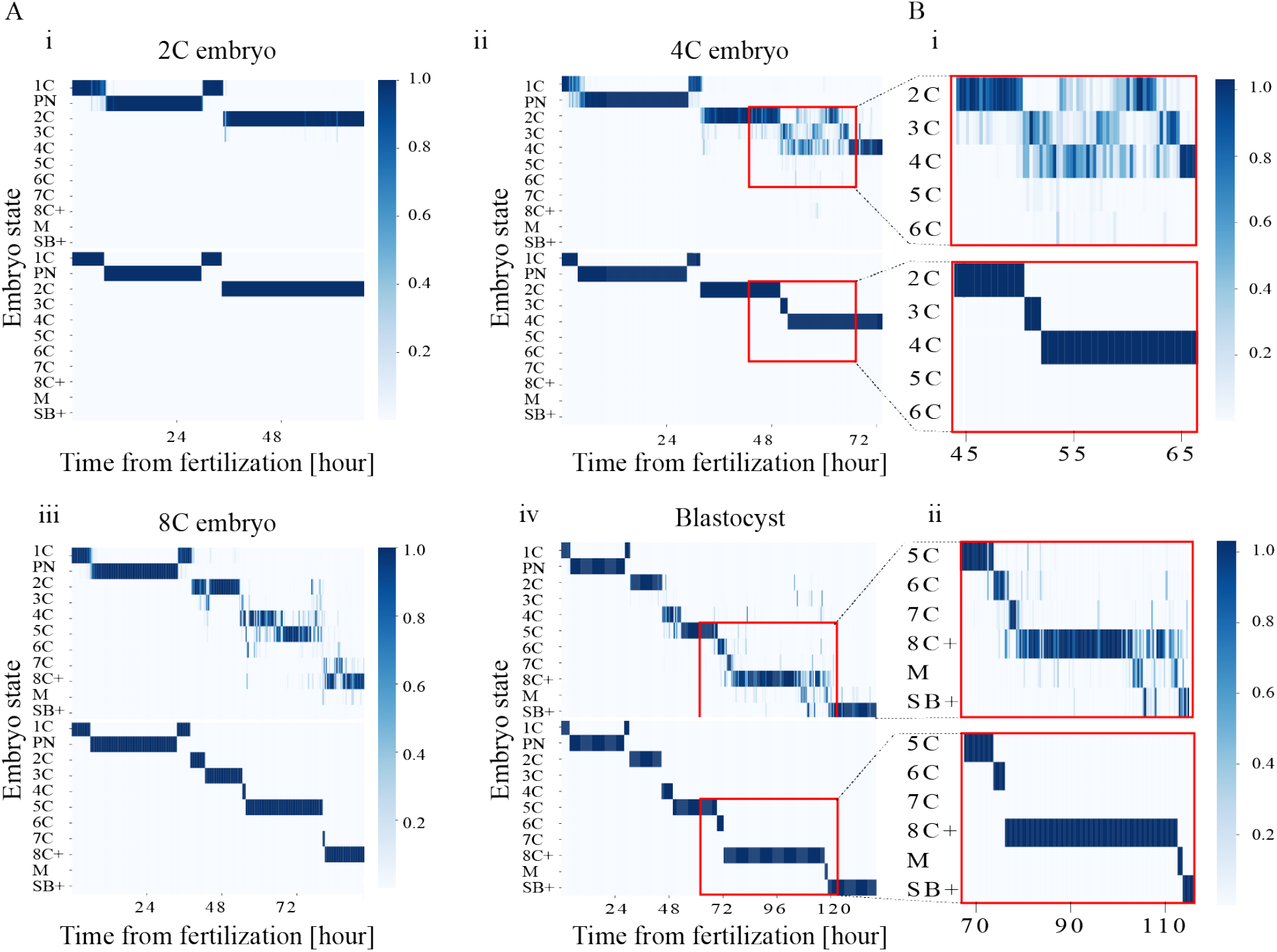
Uncertainty in the assessed embryo state probability vectors is localized to specific developmental regions. **(A)** Representative ESPM (top) and ESM (bottom) of embryos at (i) 2C, (ii) 4C, (iii) 8C, and (iv) blastocyst developmental states. (**B)** Zoom-in into the developmental regions of high uncertainty of (i) a 4C embryo, and (ii) a blastocyst. 2C: Two-cells. 4C: Four-cells. 8C: Eight-cells.

**Fig 5.**
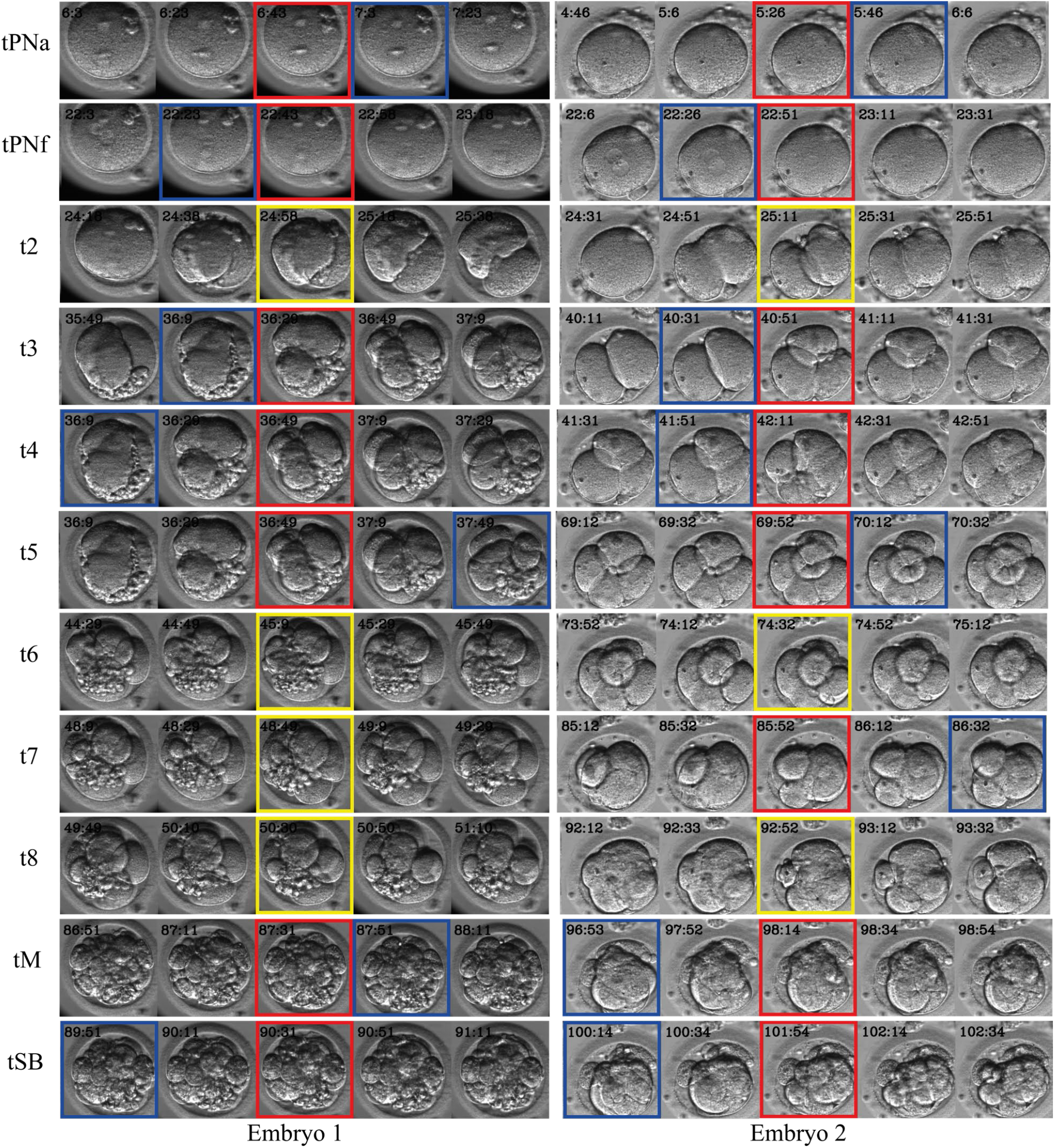
Automatic morphokinetic annotation. A comparison between the automatic morphokinetic annotations and the manual ground truth is demonstrated for two representative embryos (left and right columns). Five consecutive frame strips (rows) are centered around each ground truth event (central column). Manually-annotated frames are marked by red frames. Automatically-annotated frames are marked by blue frames. Manually and automatically co-annotated frames are marked by yellow frames. Time stamps are specified at top-left frames’ corners ([HH:MM]; top-left).

### 3.2. Classification model performance

To estimate the accuracy in the classification of the embryo states, we calculated the confusion matrix between the ESM and the manually annotated ground truth as averaged across 66,2021 frames of 1918 test set embryos (Fig 6A). Indeed, most embryo states were inferred in agreement with the ground truth, with 93% precision and 93% recall, whereas disagreements were localized to developmentally-adjacent states. Next, we plot the predicted versus the manually annotated morphokinetic events of the test set embryos (Fig 6B). Consistent with the classification accuracy of the embryo developmental states, we find an almost perfect correlation with 0.994 R-square. The average and standard deviation values of the temporal differences between the automatically and manually annotated events are summarized in Table-3. The mean error in the prediction of the pronuclei events (tPNa-tPNf) and the cleavage events (t2-t8^+^) was smaller or comparable with the time-lapse interval (20 min), which sets the lower bound of prediction accuracy (t8^+^ error: 25 min). However, tM (62 min) and tSB (42 min) mean errors spanned three time intervals or less owing to the inherent ambiguity in the definition of the exact time point of morula compaction and start-of-blastulation, respectively. Quantile-wise error distributions are shown in Table-4.

**Fig 6.**
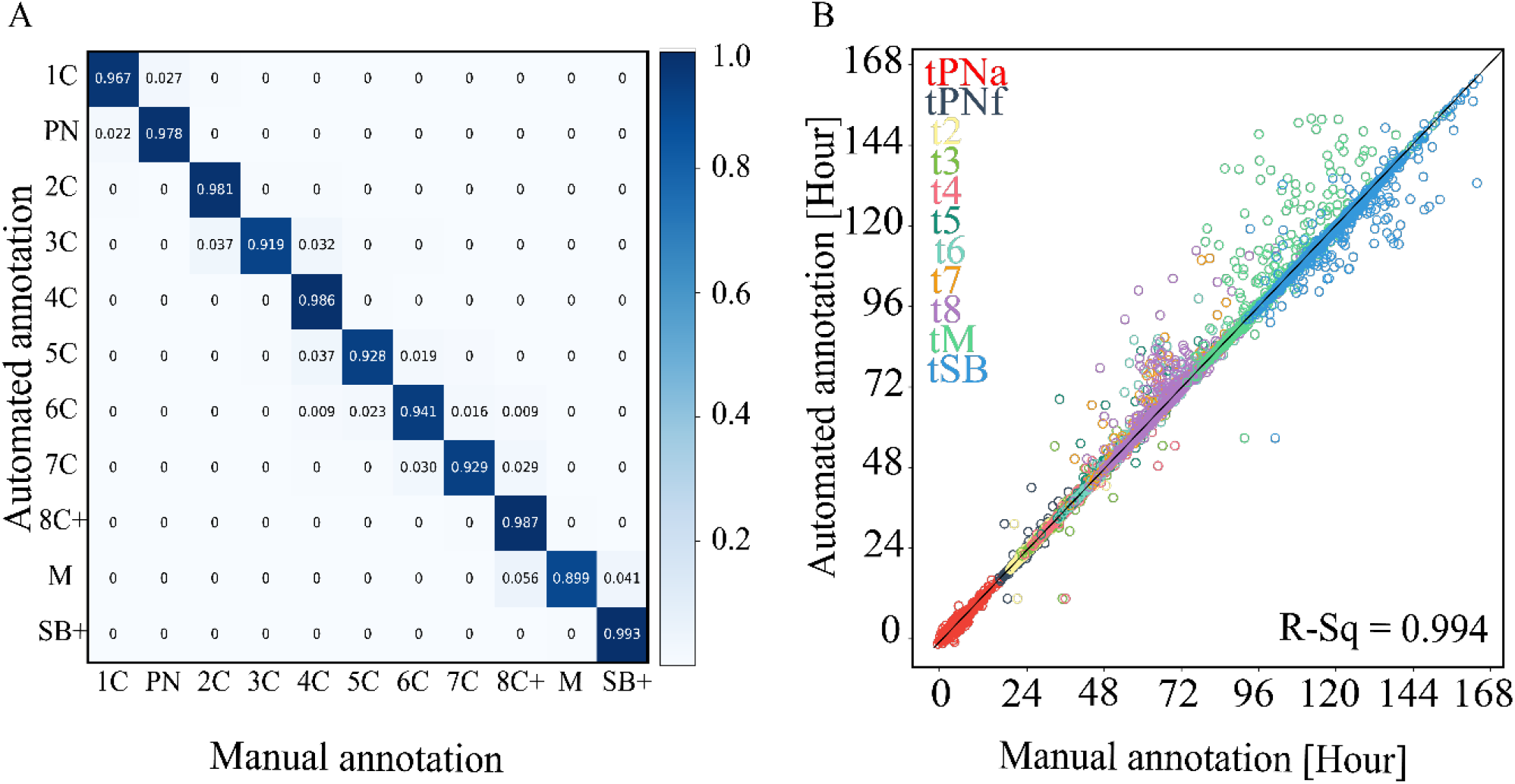
Accuracy evaluation of automatic annotation. **(A**) A confusion matrix presenting the normalized associations between manually-annotated (ground truth) and automatically-annotated (inference) test set embryo frames. For example, 3.7%, 92.8% and 0.019% of the automatically-annotated 5C frames were manually-annotated as 4C, 5C and 6C, respectively. (**B)** The automated prediction of the morphokinetic events correlate with the manually annotated ground truth with R-square 0.994 accuracy. Statics is based on 17,077 morphokinetic events expressed by 1,918 test set embryos.

**Table-3:**
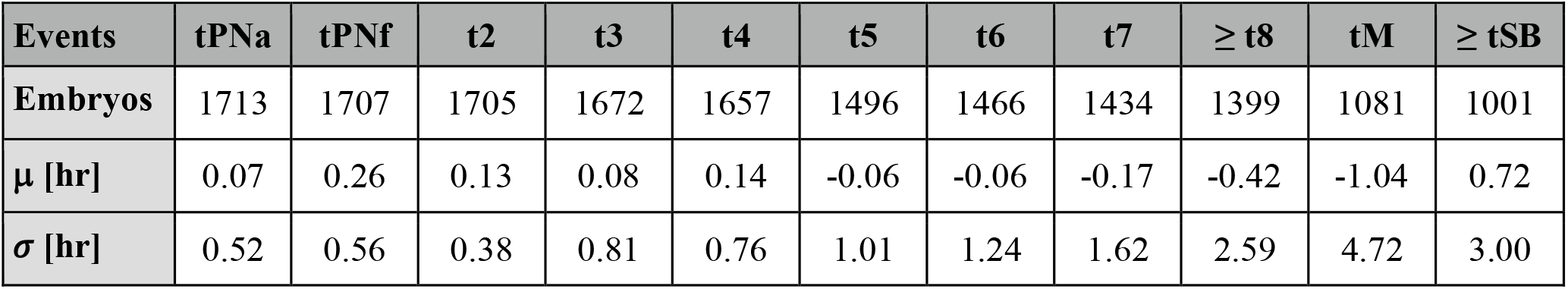
The average and standard deviation values of the temporal differences between inferred and ground truth annotations were calculated using the specified number of test set embryos. μ: Mean. *σ*: standard deviation of the mean.

**Table-4:** The distributions of the temporal differences between automatic annotation and manual ground truth were calculated for each morphokinetic event. The specified percentiles are provided in hours.

| Events | tPNa | tPNf | t2 | t3 | t4 | t5 | t6 | t7 | ≥ t8 | tM | ≥ tSB |
| --- | --- | --- | --- | --- | --- | --- | --- | --- | --- | --- | --- |
| **25%** | -0.25 | 0.25 | 0.00 | 0.00 | 0.00 | 0.00 | 0.00 | 0.00 | 0.00 | -0.25 | 0.00 |
| **50%** | 0.00 | 0.33 | 0.00 | 0.00 | 0.00 | 0.00 | 0.00 | 0.00 | 0.00 | 0.00 | 0.33 |
| **75%** | 0.33 | 0.33 | 0.33 | 0.33 | 0.33 | 0.33 | 0.33 | 0.33 | 0.33 | 0.33 | 0.66 |
| **80%** | 0.33 | 0.34 | 0.33 | 0.33 | 0.33 | 0.33 | 0.33 | 0.33 | 0.33 | 0.33 | 0.67 |
| **85%** | 0.50 | 0.34 | 0.33 | 0.33 | 0.33 | 0.33 | 0.33 | 0.33 | 0.33 | 0.33 | 1.00 |
| **90%** | 0.67 | 0.50 | 0.33 | 0.33 | 0.33 | 0.33 | 0.33 | 0.33 | 0.33 | 0.34 | 1.67 |
| **95%** | 1.00 | 0.67 | 0.34 | 0.34 | 0.34 | 0.33 | 0.37 | 0.34 | 0.67 | 0.67 | 4.67 |
| **99%** | 1.67 | 0.75 | 0.66 | 0.67 | 0.67 | 0.67 | 0.67 | 0.67 | 1.00 | 1.34 | 14.00 |

Above we presented the accuracy in the prediction of individual embryo states and morphokinetic events as we evaluated across a large dataset of embryos. However, clinical applications would also require quantitative assessment of the annotation accuracy of the morphokinetic profiles from tPNa to tSB of individual embryos, which will allow consideration of classification generality. To this end, we defined the normalized absolute temporal difference (NATD) between the automated and manual annotations as:

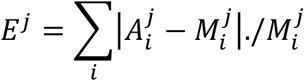

where 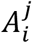 and 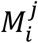 are the vectors of automatically and manually annotated morphokinetic events, denoted by index *i*, of embryo *j*, respectively. Since differences in the IVF protocols may vary between medical centers and since maternal age is linked with temporal morphokinetic profiles,^31,32^ we calculated the NATD histograms as evaluated for test set embryos stratified by clinic (Fig 7A) and by maternal age (Fig 7B). The distributions of the basal sampling error per embryo were evaluated assuming one interval difference between 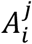 and 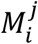 (∼18 min). Satisfyingly, we find that the NATD error distributions were broader yet statically-significantly smaller than the basal sampling error. In summary, we report unpresented accuracy statistics of automatic morphokinetic annotation ranging from single-frame embryo state prediction (Fig 6A), to inference of population-level morphokinetic events (Fig 6B, and Tables 3 and 4) and whole-embryo morphokinetic profiles (Fig 7). In addition, we demonstrate that automatic morphokinetic annotation is robust to medical center and maternal age, thus satisfying generality (Fig 7).

**Fig 7.**
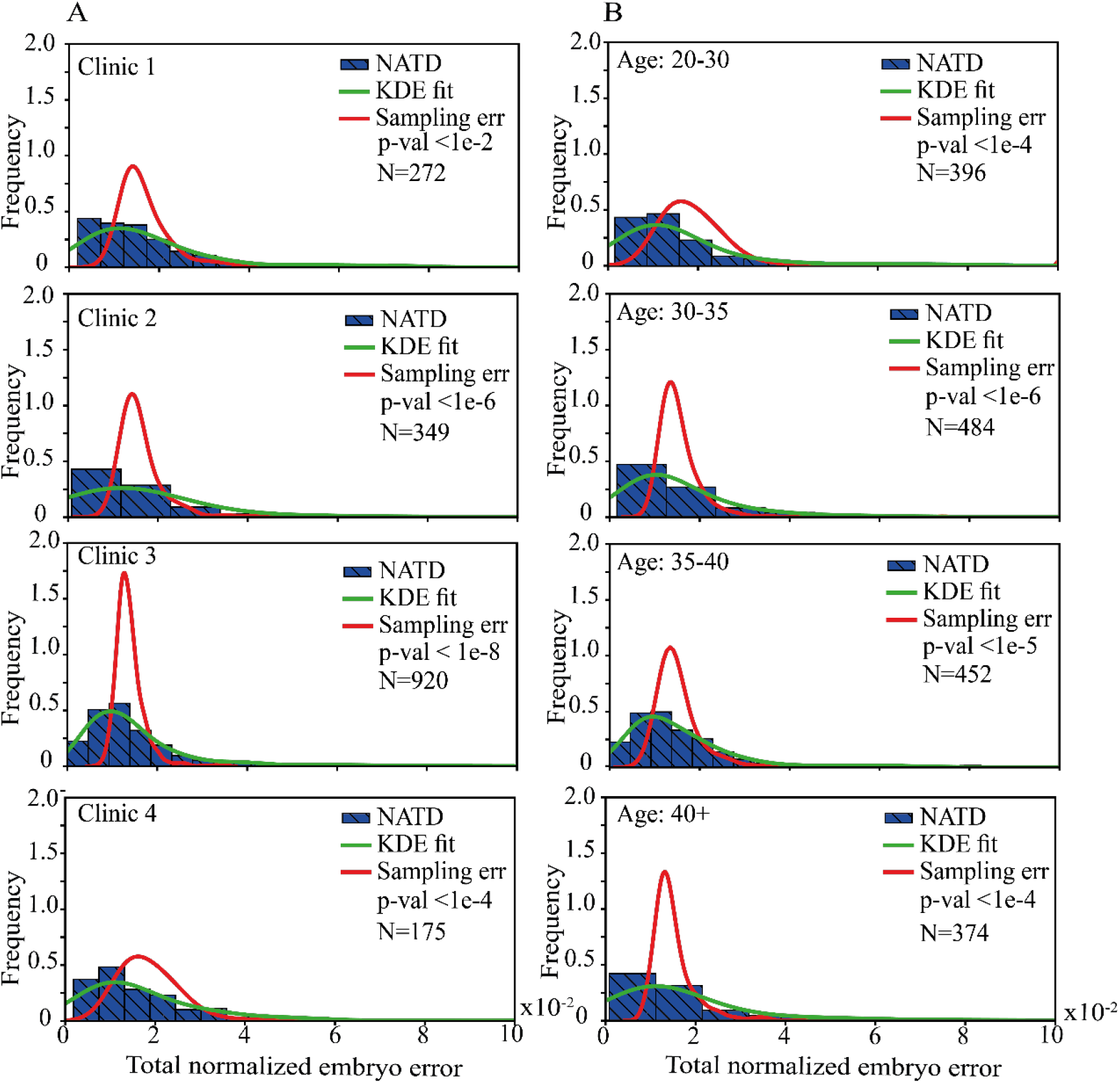
Error analysis of automatic morphokinetic annotation of embryos. The NATD histograms provide quantification of the morphokinetic difference per embryo between automatic and manual annotations. Histograms and KDE fits are plotted for each data-providing clinic (A) and across maternal age groups (B). The basal sampling error distributions account for one time-lapse interval errors per event. The number of embryos in each cohort is specified. Student’s t-test p-values are calculated between the KDE fits and the basal sampling error. NATD: Normalized absolute temporal differences. KDE: Kernel density estimation.

### 3.3. “Big data” analysis of preimplantation embryo development

Automatic morphokinetic annotation provides means for analyzing preimplantation development of a large number of embryos to provide statistical characterization, which would have been practically impossible otherwise. To address all preimplantation stages, we annotated 24,644 embryos that have been cultured inside time-lapse incubators for 120 hours or more. The generality of our analysis is based on the fact that each of such time-lapse incubator culture plates includes multiple embryos of different developmental potential with no apriority bias of maternal age, clinic, or number of retrieved oocytes per collection cycle. At each hour from time of fertilization to 120 hours, we evaluated the distribution of embryo states and calculated the 25^th^, 50^th^, 75^th^ and 95^th^ percentiles of preimplantation development with respect to the discrete metric of the twelve embryo states from FO to BL (Fig 8A). The 25^th^ percentile is defined by the embryos with the slowest dynamics, which became arrested prior to morula compaction. In comparison, the 50^th^, 75^th^ and 95^th^ percentile dynamics reach tM by end of Day-4 (96 hours), late Day-3 (68 hours), and early Day-3 (51 hours), respectively, thus demonstrating the temporal variation between embryos. Complementary to the embryo state dynamics, we calculated the temporal distributions of the morphokinetic events, the morphokinetic cell-cycle intervals, and the morphokinetic cell-synchronization intervals, as defined by the transitions between states (Fig 8B-i,ii).^33^ Here, the number of embryos that reached each morphokinetic event decreased with preimplantation development due to prior-arrest of some of the embryos. The temporal dispersion of the morphokinetic events across the embryos, as evaluated by the coefficient of variation, was significant, indicating the inherent heterogeneity between embryos of similar genetic background (denoted in Fig 8B-i,ii). Importantly, these temporal variations decreased with preimplantation development, thus demonstrating the developmental convergence of non-arrested embryos.

**Fig 8.**
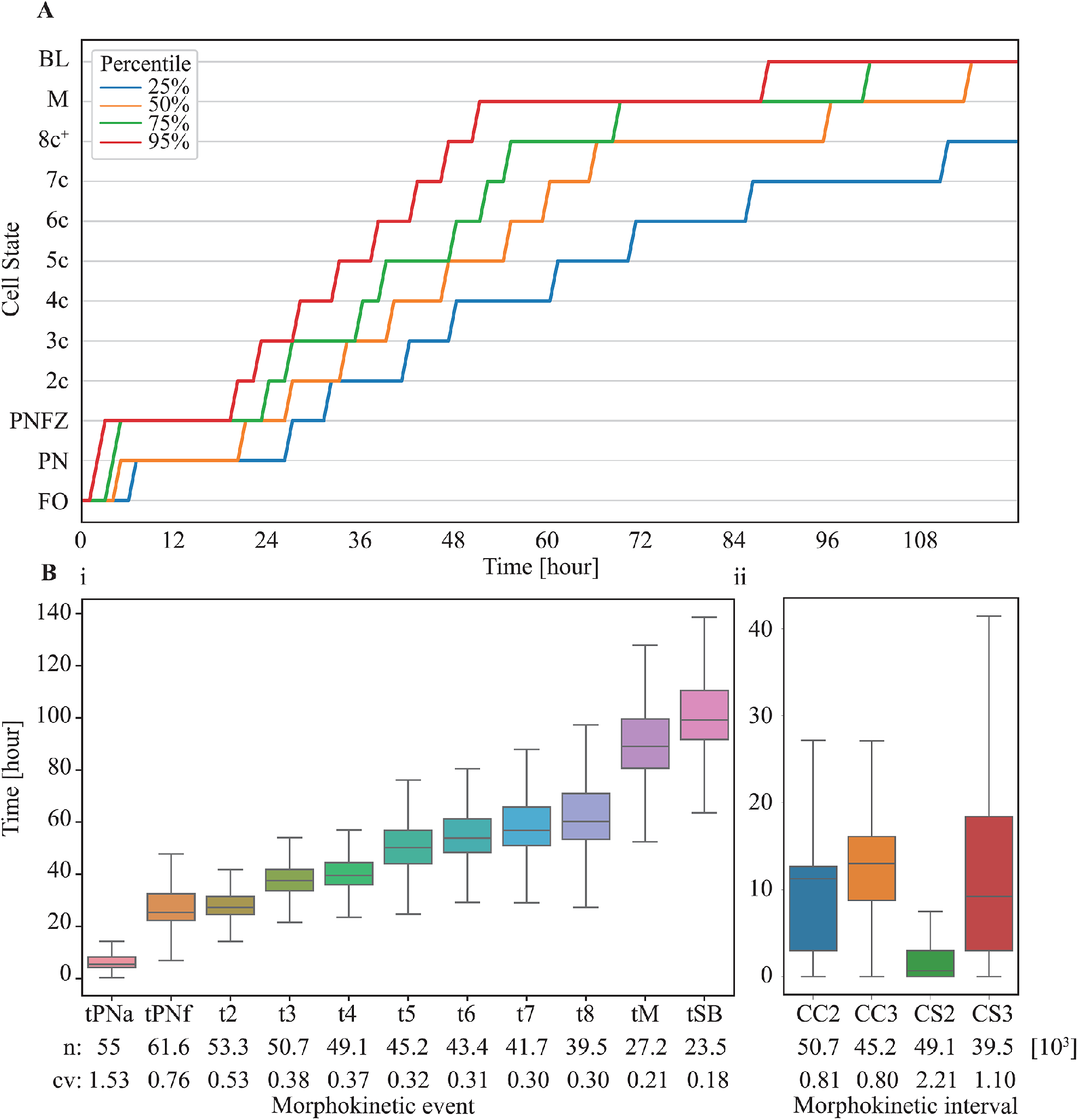
Statistics of preimplantation development of *tens of thousands* of embryos. **(A)** Embryo state trajectories are evaluated at one-hour resolution from time-of-fertilization to 120 hours as depicted for the specified percentiles across 24,644 embryos with 120 hour or longer time-lapse recordings. Statistics includes transferred and non-transferred embryos. (**B)** The temporal distributions were calculated for the specified (i) morphokinetic events and (ii) the CC and CS intervals. The number of embryos and the CV values are specified for each event below the graphs. Analyses are based on automatic inference of embryo developmental states and morphokinetic events. Whiskers depict the 5^th^, 25^th^, 50^th^, 75^th^ and 95^th^ percentiles. CC: Cell cycle. CS: Cell synchronization. CC2 = t3-t2. CC3 = t5-t3. CS2 = t4-t3. CS3 = t8-t5. CV: Coefficient of variation.

The address embryo-to-embryo variation, we performed unsupervised K-means clustering of the morphokinetic profiles. Since low-quality embryos have negligible clinical significance, we included only the high-quality embryos that are generally considered for transfer by excluding the ones that failed to reach 8C by 66 hours from fertilization (8C^-^ embryos). We clustered the embryos into K=9 cohorts, thus allowing high variation while limiting the number of clusters (Fig 9A). The clusters C0 to C8 are sorted from early to late tPNa (Fig 9B). The size of clusters C0 to C8 is listed in Table-5 and compared with the cohort of 8C^-^ low-quality embryos. Importantly, we find that the differences in preimplantation dynamics between the clusters are not associated with differences in maternal age (Fig 9C) nor in the rate of blastulation (Fig 9D).

**Fig 9.**
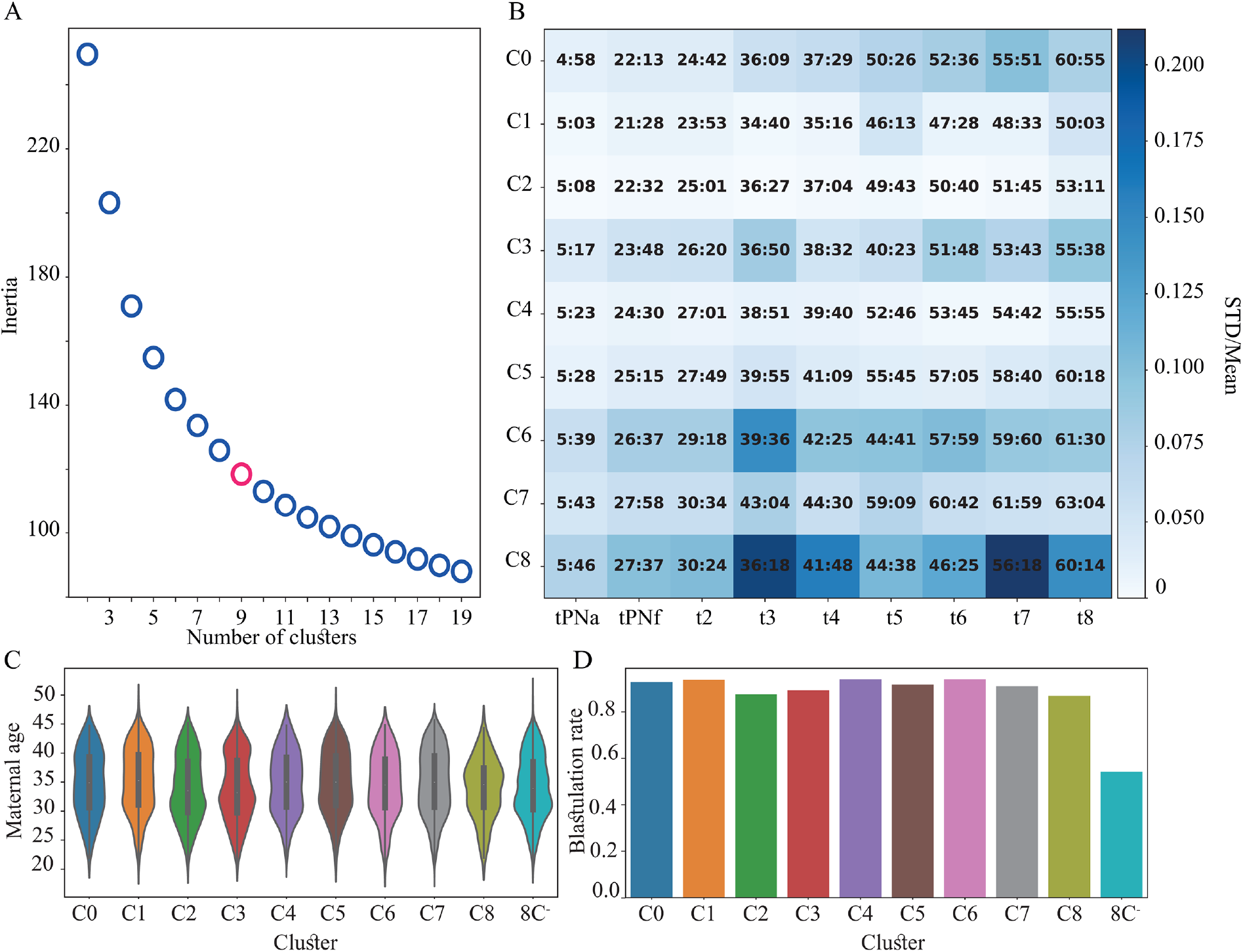
Unsupervised clustering of the embryos. **(A)** An elbow plot presenting the decrease in variance (inertia) with increasing number of clusters for the unsupervised K-means clustering of high-quality 8C^+^ embryos as performed based on the tPNa-to-t8 morphokinetic profiles. K=9 clusters was chosen. (**B)** The clusters C0 to C8 are sorted from fast (C0) to slow (C8) tPNa. (**C)** The maternal age distributions are indistinguishable across embryo clusters. (**D)** The C0-to-C8 clusters of 8C^+^ embryos reach start-of-blastulation rate, which is twofold higher than 8C^-^ embryos. Blastulation rates were calculated for embryos in each cluster that were cultured for 120 hours or longer. 8C^+/-^: embryos that reached/failed to reach 8-cells stage by 66 hours from fertilization.

**Table-5:**
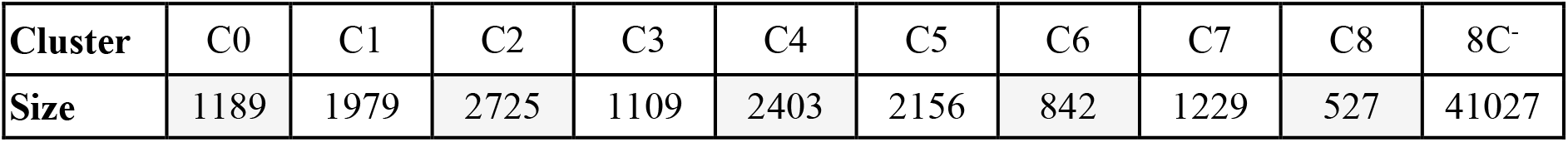
Number of embryos in each cluster. The excluded embryos that failed to reach the 8C stage within 66 hours from fertilization (8C^-^) are pooled together.

Clustering analysis reveals distinctive morphokinetic patterns that are characteristic of embryo subtypes (Fig 10A). To characterize preimplantation dynamics, we consider the first, second, and third embryo cleavage blocks (Fig 10B). All clusters maintain their order from fast to slow during the first blastomere cleavage round (PN to 2C). However, C0 embryos entered the first blastomere cleavage round early but completed the third blastomere cleavage round late. Next, we calculated the second (CS2 = t4-t3) and third (CS3 = t8-t5) cell synchronization intervals, which quantify the degree of meiotic synchronization between the blastomeres in the corresponding cleavage blocks (Fig 10C). C3, C6 and C8 embryos are the least synchronized. With respect to the third meiotic cycle, the synchrony of these clusters is as poor as the low quality 8C^-^ embryos.

**Fig 10.**
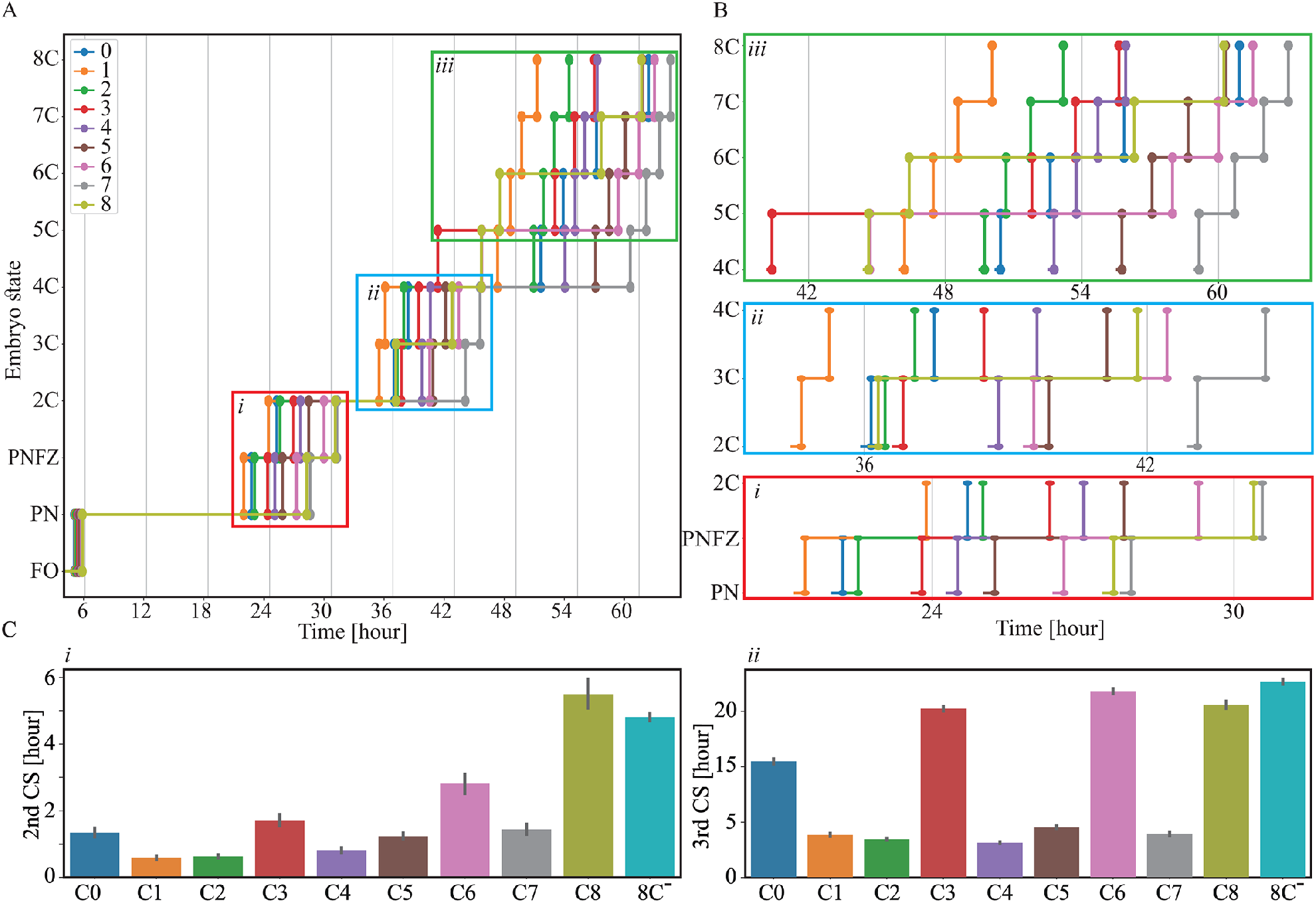
Embryo clusters are characterized by distinctive morphokinetic dynamics. **(A)** Contours of the average morphokinetic events are shown for clusters C0 to C8 and compared with the cohort of low quality 8C^-^ embryos. (**B)** Zoom-in diagrams of the first (i: PN to 2C), second (ii: 2C to 4C) and third (iii: 4C to 8C) blastomere cleavage blocks differentiate between developmentally slow and fast preimplantation dynamics. (**C)** The average cell synchronization intervals of (i) the second blastomere cleavage round (CS2 = t4-t3) and (ii) the third blastomere cleavage round (CS3 = t8-t5) are shown for C0 to C8 clusters and compared with 8C^-^ embryos. Error bars depict standard deviation.

### 3.4. The developmental potential of the embryo clusters

The rate of embryo transfer into the uterus reflects the developmental quality of the embryos as estimated by the clinicians. To assess the relationship to the developmental potential of the embryos, we plot the average implantation-versus-transfer rates of the clusters of Day-3 (Fig 11A), Day-4 (Fig 11B) and Day-5 (Fig 11C) transferred embryos, and include 8C^-^ low-quality embryos. As expected, the implantation rates of Day-5 transferred embryos are highest and of Day-3 transferred embryos are lowest. The decrease in embryo transfer rates between Day-3 and Day-5 transfers is due to the decrease in the number of transferred blastocysts per cycle. Despite the fact that the cluster distributions of the available embryos per oocyte collection cycle are not specified, the average implantation rates are positively correlated with the transfer rates, thus conforming the capacity of morphokinetic profiling in predicting embryo quality. C1 and C2 clusters consistently show highest implantation rate and high transfer rates. C3, C6 and C8 clusters have the lowest implantation rates and low transfer rates during embryo cleavage (Day-3) and morula compaction (Day-4) stages, which is consistent with the poor cell cleavage synchronization of these embryos during the third meiotic cycle (Fig 10C). Hence, our unsupervised clustering analysis confirms the role of CS3 as a marker of embryo developmental potential.^21,34^ We note that blastocyst selection for transfer on Day-5 significantly improves the implantation rates of C3 and C6 clusters (C8 embryos have zero implantation rate).

**Fig 11.**
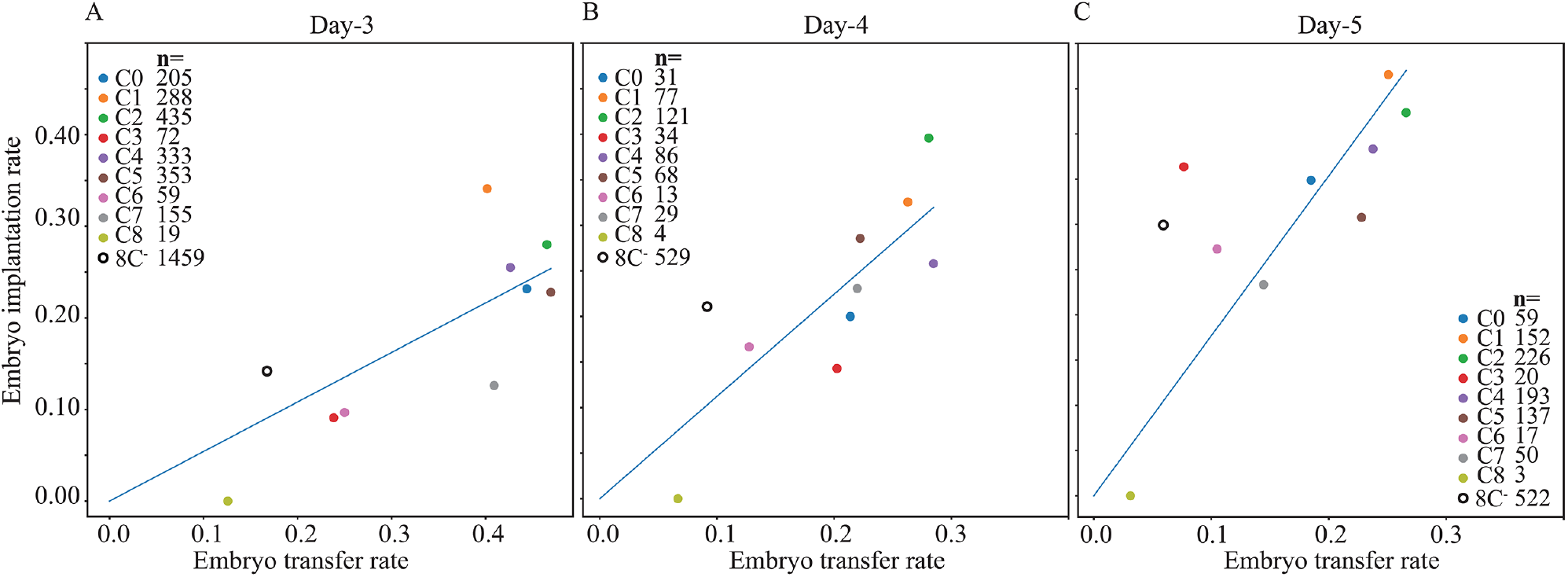
Transfer and implantation rates across embryo clusters. The average implantation rates are plotted as a function of the average implantation rates of clusters C0 to C8 and 8C^-^ embryos (at 66 hours from fertilization). Implantation-versus-transfer rates are plotted for (A) Day-3 cleavage stage embryo transfers (66 to 74 hours), (B) Day-4 embryo transfers (74 to 110 hours), and (C) Day-5 blastocyst transfers (110 to 125 hours). The number of transferred embryos is specified in the legends.

## 4. Discussion

Temporal profiling of the morphokinetic events has been demonstrated to support the evaluation of the developmental potential of embryos and improve implantation rates by allowing the selection of the highest quality embryos for transfer.^35^ Computationally, morphokinetic annotation effectively reduces the dimensionality of a video representation of the embryos from ∼100 Mb to ∼100 bytes, thus preventing overfitting and improving accuracy given a finite dataset and computational power. However, the medical utilization of the available morphokinetic classification algorithms in IVF treatments is hindered by the time-consumption that is required by manual annotation and the intra- and inter-observer variations. Hence, there is a substantial need for algorithms that perform automatic morphokinetic annotation of the embryos’ video files that are continuously recorded by time-lapse incubators.

To date, automatic annotation of blastomere cleavage events was reported using different machine learning methodologies.^36-38^ Recently, automatic annotation of the entire course of preimplantation development from pronuclei appearance to blastulation was reported, which can potentially support embryo selection for Day-5 transfers. Using image processing tools, Feyeux et al. report 92% accuracy, which is likely insufficient for clinical applications as a stand-alone automatic decision-support tool.^39^ Using an expansive retrospective dataset to train a CNN classifier, we assess the probabilities of the embryo stages to appear in each frame, and employ monotonic regression to extract the time points of the morphokinetic events of the embryos from pronuclei appearance to start-of-blastulation. In this manner, we generate an automatic tool for performing morphokinetic annotation with unpresented accuracy (R-square 0.994), whose error distribution is largely generated by the discrete nature of time-lapse imaging. The fact that similar error distributions are obtained by each data-providing clinic and that are independent of maternal age demonstrates generality. Hence, the algorithm that we present here has the capacity to mediate the clinical integration of embryo selection based on morphokinetic evaluation of the developmental potential in IVF settings.

Using automated annotation, we calculated the temporal distributions of the morphokinetic profiles of 24,644 embryos that were cultured for > 120 hours. We found a large variation among embryos that implies the existence of a collection of embryo subtypes. To exclude statistical contributions of low-quality embryos, we restricted our analysis to high-quality embryos (8C^+^ at 66 hours from fertilization) that serve as valid candidates for transfer. Using K-means unsupervised clustering of 14,159 8C^+^ embryos, we defined nine cohorts of embryos that are characterized by distinctive morphokinetic dynamics. This included “fast” and “slow” developing embryos, as well as embryos that “start fast” and “finish slow”. The maternal age distributions of the clusters overlapped and were statistically indistinguishable from the low quality 8C^-^ embryos. This supports the assumption that maternal age affects the size of the oocyte collection cycle, however the embryos in each OCC from young and older women are heterogeneous.^40^ Unlike the blastulation rate, the implantation rates the embryo clusters is different. This reflects the computational challenge of elucidating the variation among high-quality 8C^+^ embryos excluding low quality embryos. We found a positive correlation between the average implantation and transfer rates of the embryo clusters, indicative of the utility of morphokinetic annotation in predicting embryo developmental potential. We identified three clusters that were characterized by low implantation rates. Importantly, these embryo clusters were distinctively marked by poor synchronization in the third meiotic cell cleavage cycle. The embryo clustering analysis that we define here provides a computational framework for researchers and clinicians to assess the variation among the embryos in each oocyte collection cycle, thus supporting the multi-step embryo selection decision-making strategy.

## Data Availability

The copyrights of the code are owned by Yissum?the technology transfer company of The Hebrew University of Jerusalem. Requests can be sent to A.B. The clinical data are owned by Hadassah Medical Center and by Clalit Health Services. Restrictions apply to the availability of these data, which were used anonymously under ethical agreements with each clinic separately for this study, and so are not made publically available. Access requests can be directed to A.B.M. (Hadassah Medical Center), Y.O. (Kaplan Medical Center), I.H.V (Soroka University Medical Center), Y.S. (Women's Hospital, Rabin Medical Center).

## Ethical approval

This research was approved by the Investigation Review Boards of the data-providing medical centers: Hadassah Hebrew University Medical center IRB number HMO 558-14; Kaplan Medical Center IRB 0040-16-KMC; Soroka Medical Center IRB 0328-17-SOR; Rabin Medical Center IRB 0767-15-RMC.

## Code and data availability

The copyrights of the code are owned by Yissum–the technology transfer company of The Hebrew University of Jerusalem. Requests can be sent to A.B. The clinical data are owned by Hadassah Medical Center and by Clalit Health Services. Restrictions apply to the availability of these data, which were used anonymously under ethical agreements with each clinic separately for this study, and so are not made publically available. Access requests can be directed to A.B.M. (Hadassah Medical Center), Y.O. (Kaplan Medical Center), I.H.V (Soroka University Medical Center), Y.S. (Women’s Hospital, Rabin Medical Center).

